# High Levels of Co-detection of Norovirus and Other Enteric Pathogens in Hospitalized Patients with and without Acute Gastroenteritis in Bangladesh

**DOI:** 10.1101/2025.10.20.25338336

**Authors:** Farzana Fariha, Adiba Hassan, Sharmeen Akter Urmi, Mohammad Enayet Hossain, Probir Kumar Ghosh, Md. Mahfuzur Rahman, Faruq Abdullah, Kamal Ibne Amin Chowdhury, Zarin Abdullah, Rashi Gautam, Sayera Banu, Jan Vinjé, Sara A. Mirza, Mustafizur Rahman, Paul A. Gastañaduy, Umesh D Parashar, Syed M Satter

## Abstract

**Background:** The contribution of various enteropathogens to the occurrence of acute gastroenteritis (AGE) remains uncertain in highly endemic settings. We describe the frequency of norovirus-only detections and norovirus co-detections in hospitalized patients in Bangladesh and compare their clinical severity and viral load.

**Methods:** From March 2018–October 2021, 1,250 AGE cases and 1,250 non-AGE controls of all ages were enrolled at 10 tertiary care hospitals in Bangladesh. All norovirus-positive AGE cases (n=111) and non-AGE controls (n=182), and a randomly selected subset of 126 norovirus-negative AGE cases, were tested for other enteric viral, bacterial, and parasitic co-pathogens with quantitative real-time PCR assays. We used cycle threshold (Ct)-values as a proxy for viral load and the Vesikari scale to assess disease severity.

**Results:** Overall, 92% (218/237) of AGE cases had ≥1 enteropathogen detected. Among 293 norovirus-positive AGE cases and non-AGE controls, 88 (30%) were norovirus-only detections and 205 (70%) were norovirus co-detections. Norovirus-rotavirus was the predominant co-detection, found in 140 (68%) of 205 norovirus co-detections. No differences in clinical severity were observed among AGE cases with norovirus-only versus norovirus co-detections. The median (interquartile range) Ct-values among genogroup II norovirus-only AGE cases, norovirus co-detection AGE cases, and norovirus-positive non-AGE controls were 26 (21–31), 25 (20–29), and 25 (21–30), respectively.

**Conclusions:** The frequent co-detection of norovirus with other enteropathogens, especially rotavirus, along with overlapping Ct-values in patients with and without AGE, and between norovirus-only and norovirus co-detections, complicates attributing norovirus as a primary cause of AGE in hospitalized patients in Bangladesh.

---

The etiological diagnosis of acute gastroenteritis (AGE) is essential to prioritizing preventive interventions, including vaccines. Most often, epidemiological studies assessing the causes of AGE focus on identifying single enteric pathogens among symptomatic individuals, with few studies concomitantly assessing the role of other enteric pathogens or comparing detection rates among persons with and without diarrhea. In a recent study in Bangladesh, we found a higher prevalence of norovirus among hospitalized controls with no AGE symptoms compared to hospitalized cases with AGE symptoms (15% and 9%, respectively) [5], which suggested a lack of association between norovirus and diarrhea. However, the etiologic role of norovirus as a cause of diarrhea is well established.[6–9] A higher prevalence of norovirus in controls compared to cases is often attributed to asymptomatic infections and prolonged shedding following recovery from illness. This is expected to be more common in endemic settings with high levels of exposure.[6, 10] Further complicating the picture were overlapping cycle threshold (Ct)-values, a proxy for viral load and clinical disease, and high levels of co-infection with other enteric pathogens, among norovirus-positive cases and controls.

Building on the prior study, we leveraged the expanded testing done for a range of enteric viral, bacterial, and parasitic pathogens to compare the detection rates of these pathogens among norovirus-positive hospitalized AGE cases and non-AGE controls. We aimed to (1) summarize the frequency of co-detections with norovirus and other enteric pathogens in hospitalized patients; (2) compare the clinical severity of AGE cases with single norovirus detections versus norovirus co-detections; and (3) assess for differences in Ct-values between AGE cases with norovirus-only detection compared to AGE cases with norovirus co-detections and non-AGE controls.

## Methods

### Data, population, and testing

Participants were enrolled from 10 tertiary care governmental and non-governmental hospitals across seven of the eight divisions in Bangladesh from March 2018 to October 2021.[5] AGE cases were patients hospitalized for rehydration with ≥3 episodes of watery or loose stool or ≥1 episode of forceful vomiting during a 24-hour period with symptoms lasting ≤7 days. Non-AGE controls were patients hospitalized without AGE symptoms during the 14 days prior to enrollment. Controls were matched to cases by sex, age group (0–4, 5–17, 18–34, 35–64, or ≥65 years), facility, and enrollment period (within 30 days). A total of 1,250 AGE cases and 1,250 non-AGE controls were enrolled in the main study. Demographic and clinical information were collected from medical records and by standardized interviews conducted by trained physicians and field assistants upon enrollment.

This study included 293 patients (111 AGE cases and 182 non-AGE controls) who tested positive for norovirus in the main study (Figure 1). To understand the overall distribution of enteropathogens among AGE cases, we separately assessed a randomly selected subset of 126 norovirus-negative AGE cases. Stool samples were collected from all patients at enrollment and tested at the International Center for Diarrheal Disease Research, Bangladesh (icddr,b) for additional enteric viral, bacterial, and parasitic co-pathogens with quantitative real-time PCR assays.

**Figure 1:**
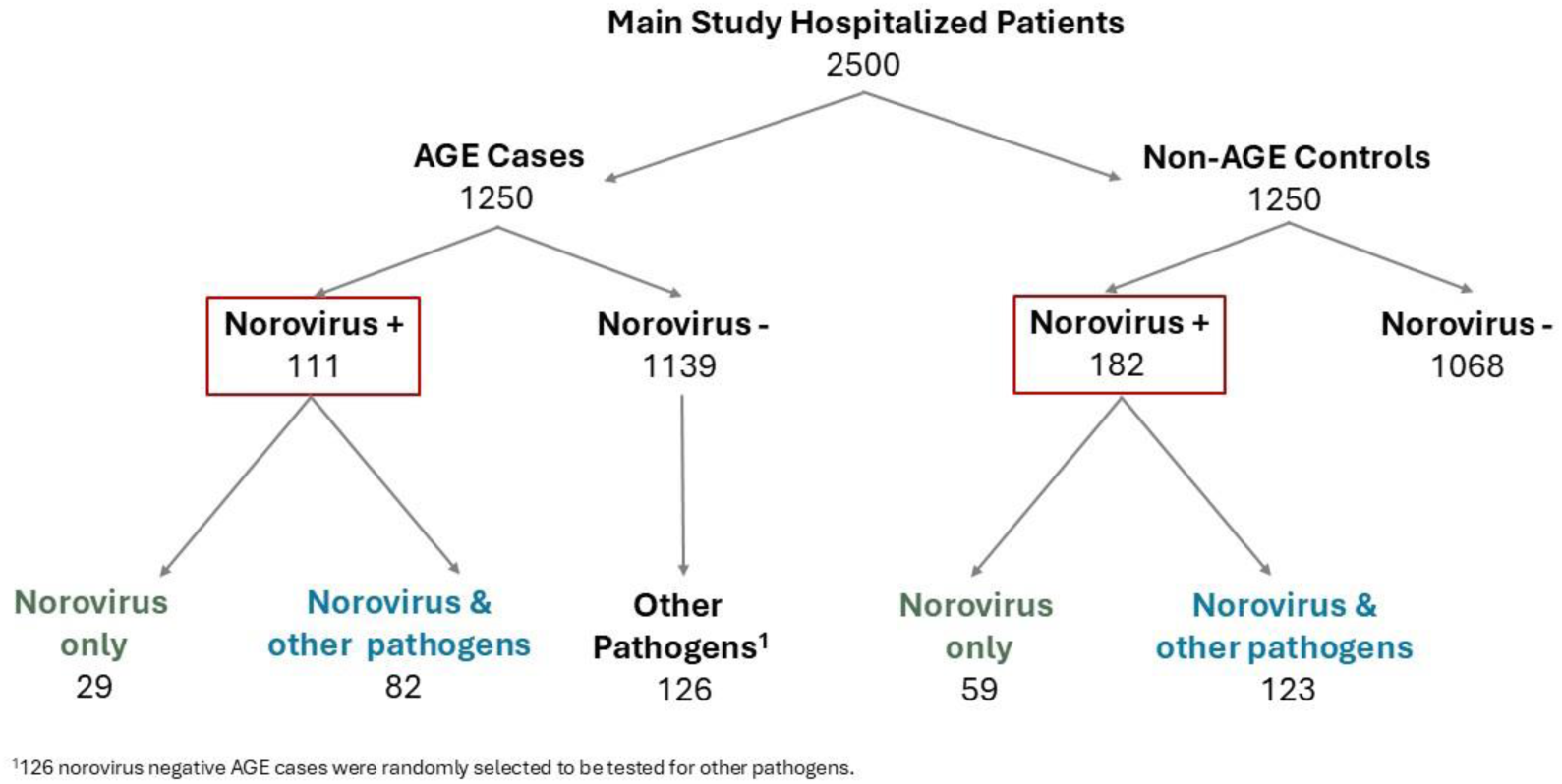
Flow chart of hospitalized patients selected from the main study. This study included norovirus-positive patients only (red box), of whom 111 were acute gastroenteritis (AGE) cases and 182 non-AGE controls in the main study. Norovirus-positive patients were further categorized into those who only had norovirus detected vs. norovirus and other pathogen co-detected.

### Nucleic Acid Extraction and Detection of Enteropathogens

Total nucleic acid was extracted from 200 µL of diluted stool supernatant using the Chemagic viral NA/gDNA kit (PerkinElmer, MA, USA), as per the manufacturer’s instructions performed on an automated nucleic acid extraction system, the chemagic™ 360 instrument. The nucleic acids were eluted in 100 µL of elution buffer and stored at −80°C until further use. TaqMan-based real-time RT-PCR was carried out for detection of norovirus GI and GII as described previously,[5, 11, 12] as well as for rotavirus, adenovirus, sapovirus, and astrovirus.[13–15] For rotavirus, the double-stranded RNA was denatured at 95°C for 5 minutes prior to the RT-PCR assay. Briefly, RT-qPCR assays were performed with TaqPath™ 1-Step Multiplex Master Mix kit (Thermo Fisher Scientific) in a final reaction volume of 20 μl. The master mixes consisted of 5 μl 4× TaqPath 1 step Multiplex Master Mix, 0.5 μl of each primer (20 μM), 0.5 μl probe (10 μM), 8.5 μl nuclease-free water, and 5 μl of nucleic acid. The PCR reactions were run at the following thermal cycling conditions: 25°C for 2 min in UNG incubation, 53°C for 10 min, 95°C for 2 min, followed by 45 cycles of 95°C for 15 s, and 55°C for 1 min. PCR was performed on a CFX96 Touch™ Real-time PCR Detection System (Bio-Rad Laboratories, Inc., Hercules, CA, USA).

To detect bacterial and protozoan parasites, extracted DNA was tested by FTlyo bacterial gastroenteritis diagnostic kit (Fast Track Diagnostics, Luxembourg) and FTD stool parasites kit (Fast Track Diagnostics) using dual-labelled molecular probes specific for each pathogen according to the kit recommendations.

### Classification as norovirus-only detection versus norovirus co-detection

Patients who tested positive only for norovirus were categorized as a norovirus-only detection, while patients who tested positive for norovirus and one or more enteric viruses (astrovirus, rotavirus, adenovirus, and sapovirus), enteric bacterial pathogens (*Campylobacter* spp., *Clostridium difficile*, *Salmonella* spp., *Shigella* spp., Shiga toxin-producing *Escherichia coli*, and *Yersinia enterocolitica*), or enteric parasitic pathogens (*Entamoeba histolytica*, *Cryptosporidium* spp., and *Giardia lamblia*) were categorized as a norovirus co-detection.

### Norovirus-only detections versus norovirus co-detections among AGE cases and non-AGE controls

We summarized and compared the distribution of norovirus-only detections, norovirus co-detections, specific pathogen combinations, and the number of pathogens detected (two, three, or four or more pathogens) by AGE cases and non-AGE controls. We also assessed differences in sociodemographic and clinical characteristics between norovirus-only detections and norovirus co-detections. Pearson χ^2^ or Fisher’s exact tests compared differences in the distribution of categorical variables, while Wilcoxon rank sum tests compared differences in the median of continuous variables. Statistical significance was defined as *P*<0.05.

### Differences in clinical severity of AGE cases with norovirus-only detections versus norovirus co-detections

We investigated whether co-infections with more than one enteric pathogen were associated with more severe illness compared to infections with a single pathogen. Among AGE cases only, we calculated the 20-point Vesikari scale to assess disease severity. Severity of illness was categorized as mild (score of 1–6), moderate (score of 7–10), or severe (score of ≥11).[16] Box plots of Vesikari scores were created to visually compare the severity of disease between AGE cases with norovirus-only detection, norovirus co-detections, and the most prevalent co-detections (i.e., norovirus-rotavirus and norovirus-rotavirus-adenovirus co-detections).

### Differences in Ct-values in AGE cases with norovirus-only detection versus Ct-values in AGE cases with norovirus co-detections and non-AGE controls

We expected Ct-values would be lower among AGE cases with norovirus-only detection compared to AGE cases with norovirus co-detections. Similarly, we expected Ct-values would be lower among AGE cases with norovirus-only detection compared to non-AGE controls with norovirus-only detection or norovirus co-detections. We compared the distribution of Ct-values between these groups separately by genogroup using kernel density plots.

Data management, statistical analysis, and data visualization were performed using STATA SE 18.

## Results

Among 293 norovirus-positive AGE cases and non-AGE controls included in this study, 30% (88/293) were norovirus-only detections and 70% (205/293) were positive for additional pathogens (Table 1). The proportion of co-detections among AGE cases (74%) was only slightly higher compared with non-AGE controls (68%), and not statistically significant (*P*=0.25). Viral-viral co-detections were more common than viral-bacterial or viral-parasitic co-detections. Among 205 norovirus co-detections, 86% (176/205) included at least one additional viral pathogen, with rotavirus and adenovirus found in 82% (140/176) and 47% (82/176) of these specimens, respectively. Despite the predominance of viruses, 70% (78/111) of AGE cases received antibiotics before or during hospitalization (Supplementary Table 1). Rotavirus was also predominant among norovirus-negative AGE cases, with a detection rate of 80% (101/126) (Supplementary Table 2a). Although viruses predominated in all age groups, they were more common among infants and young children aged less than 5 years, while bacteria were more common in older children and adults (Supplementary Table 2b). Applying the positivity rate for any pathogen among norovirus-negative AGE cases to untested AGE cases, we estimate that we would have identified one or more enteric pathogens in 86% (1078/1250) of hospitalized patients with AGE in our population if complete testing would have been done (Supplementary Table 2a, Figure 1).

**Table 1:**
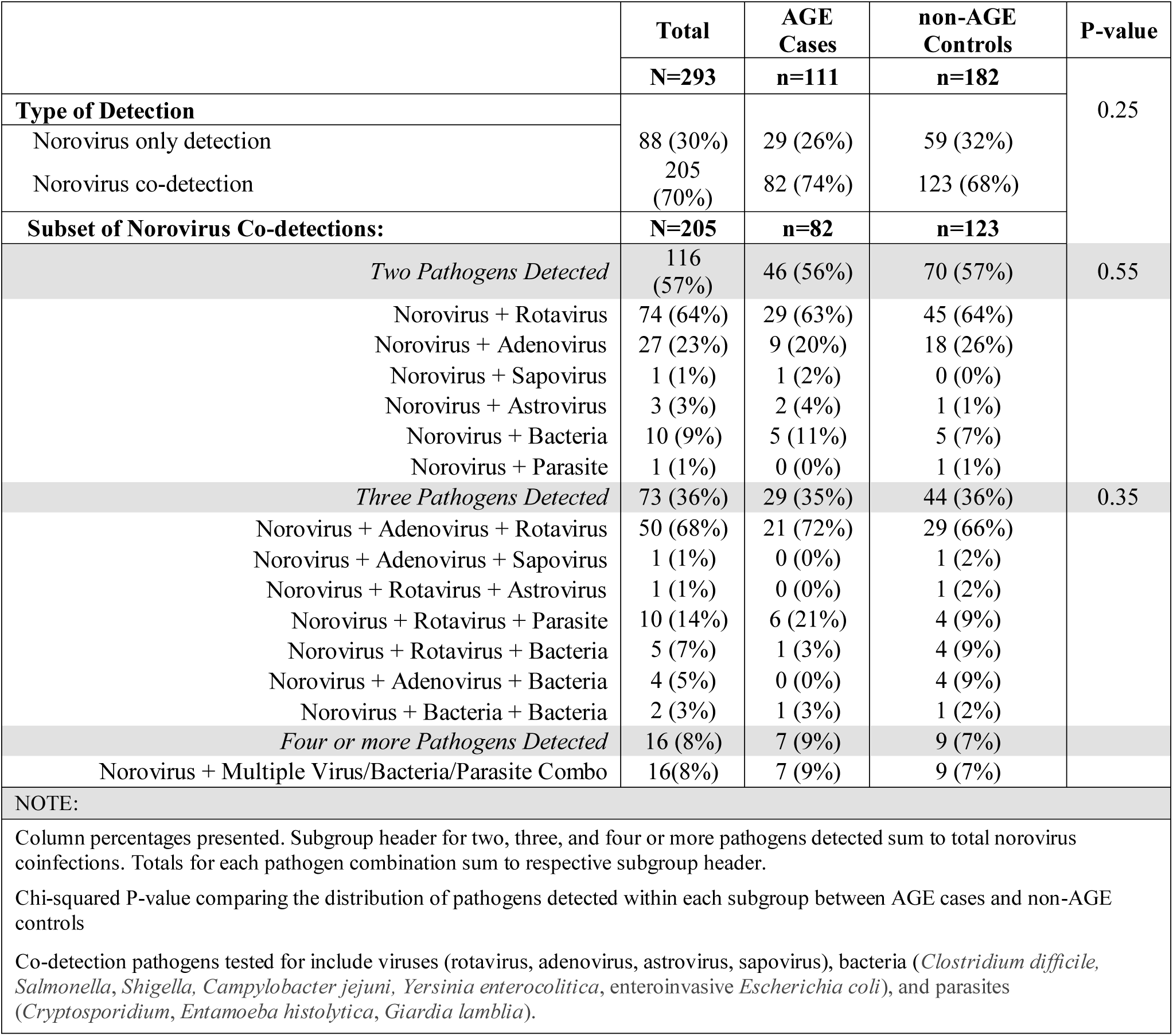
Distribution of enteric pathogens identified together with norovirus among hospitalized acute gastroenteritis (AGE) cases and non-AGE controls in Bangladesh. N=293.

Sociodemographic and clinical characteristics did not differ statistically between the norovirus-only and norovirus co-detection groups (Table 2). Based on the Vesikari scale, the majority of AGE cases (79%) were categorized as severe (Supplementary Table 1), with no differences in the proportion categorized as severe between AGE cases with norovirus-only detections and AGE cases with norovirus co-detections. Similarly, median Vesikari scores were comparable between AGE cases with norovirus-only detections and AGE cases with norovirus co-detections, including norovirus-rotavirus and norovirus-rotavirus-adenovirus co-detections (Figure 2).

**Figure 2:**
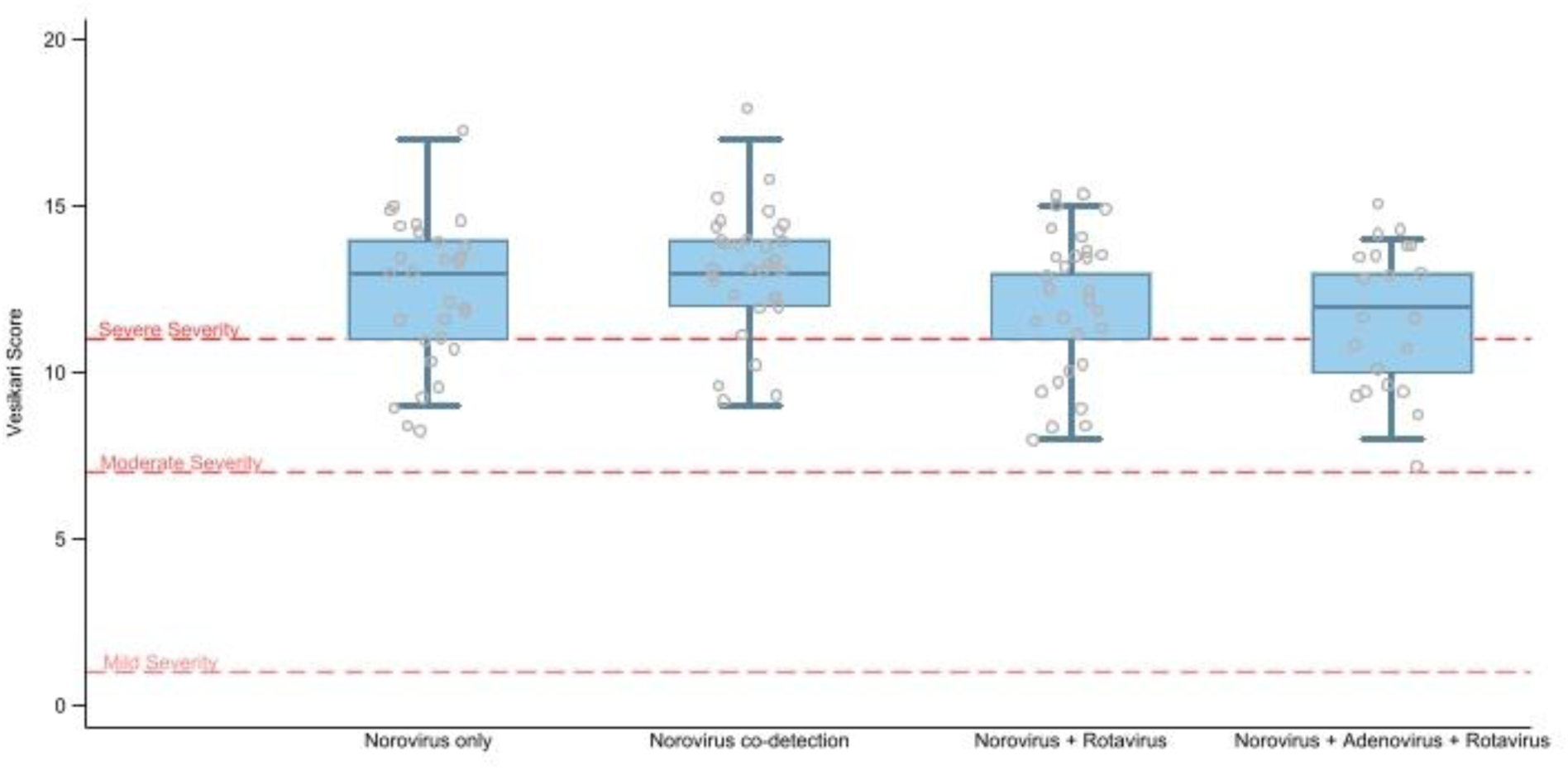
Box (median and interquartile range) and scatter (individual score) plots of Vesikari score comparing severity of acute gastroenteritis cases from 10 tertiary case hospitals in Bangladesh from March 2018-October 2021. Norovirus + rotavirus and norovirus + adenovirus + rotavirus are subsets of most prevalent (20 or more) norovirus co-detection cases.

**Table 2:** Characteristics of patients hospitalized in ten tertiary hospitals in Bangladesh from March 2018 to October 2021 by type of norovirus infection.

|  | Total | Norovirus only-detection | Norovirus co-detection | P-value |
| --- | --- | --- | --- | --- |
|  | N=293 | N=88 | N=205 |  |
| **Demographic and Socioeconomic** |  |  |  |  |
| Region |  |  |  | 0.03 |
| Barisal | 61 (21%) | 12 (14%) | 49 (24%) |  |
| Chittagong | 26 (9%) | 6 (7%) | 20 (10%) |  |
| Dhaka | 37 (13%) | 10 (11%) | 27 (13%) |  |
| Khulna | 20 (7%) | 6 (7%) | 14 (7%) |  |
| Rajshahi | 110 (38%) | 41 (47%) | 69 (34%) |  |
| Rangpur | 16 (5%) | 9 (10%) | 7 (3%) |  |
| Sylhet | 23 (8%) | 4 (5%) | 19 (9%) |  |
| Age (years), median (IQR) | 1 (0.6, 2) | 1 (0.7, 2) | 1 (0.7, 2) | 0.67 |
| Age category (years) |  |  |  | 0.47 |
| 0-4 | 245 (88%) | 69 (85%) | 176 (89%) |  |
| 5-17 | 6 (2%) | 3 (4%) | 3 (2%) |  |
| 18+ | 28 (10%) | 9 (11%) | 19 (10%) |  |
| Sex |  |  |  | 0.46 |
| Female | 91 (31%) | 30 (34%) | 61 (30%) |  |
| Male | 202 (69%) | 58 (66%) | 144 (70%) |  |
| Number of permanent household members, median (IQR) | 5 (4, 6) | 5 (4, 6) | 5 (4, 6) | 0.78 |
| Maternal education status for patients <18 years of age |  |  |  | 0.67 |
| Illiterate | 11 (4%) | 4 (5%) | 7 (4%) |  |
| Primary/PCS | 75 (29%) | 20 (27%) | 55 (30%) |  |
| Class eight/JSC | 94 (37%) | 28 (37%) | 66 (36%) |  |
| SSC/HSC | 63 (25%) | 19 (25%) | 44 (24%) |  |
| Graduate/Master and above | 9 (4%) | 4 (5%) | 5 (3%) |  |
| Religious schooling | 4 (2%) | 0 (0%) | 4 (2%) |  |
| Monthly family income (taka), median (IQR) | 15,000 (10000, 20000) | 12,000 (9000, 20500) | 15,000 (10000, 20000) | 0.43 |
| Shared bathroom or toilet with another household |  |  |  | 0.74 |
| No | 190 (65%) | 56 (64%) | 134 (66%) |  |
| Yes | 102 (35%) | 32 (36%) | 70 (34%) |  |
| **Clinical** |  |  |  |  |
| Patient transferred from another facility/hospital |  |  |  | 0.19 |
| No | 279 (95%) | 86 (98%) | 193 (94%) |  |
| Yes | 14 (5%) | 2 (2%) | 12 (6%) |  |
| Contact in last 10 days before illness with someone with diarrhoea or vomiting |  |  |  | 0.85 |
| No | 264 (90%) | 80 (91%) | 184 (90%) |  |
| Yes | 28 (10%) | 8 (9%) | 20 (10%) |  |
| Duration of hospital stay (days), median (IQR) | 3 (2, 7) | 4 (2, 8) | 3 (2, 6) | 0.16 |
| Vesikari score |  |  |  | 1.00 |
| Moderate (7-10) | 23 (21%) | 6 (21%) | 17 (21%) |  |
| Severe ($\geq 11$) | 88 (79%) | 23 (79%) | 65 (79%) | |
| Antibiotic use before or during hospitalization prior to stool collection |  |  |  | 0.30 |
| No | 82 (28%) | 21 (24%) | 61 (30%) |  |
| Yes | 211 (72%) | 67 (76%) | 144 (70%) |  |
| Antibiotic type |  |  |  | 0.079 |
| Oral (outside of hospital) | 12 (4%) | 2 (2%) | 10 (5%) |  |
| Injectable (outside of hospital) | 2 (1%) | 2 (2%) | 0 (0%) |  |
| Given at hospital | 197 (67%) | 63 (72%) | 134 (65%) |  |
| Did not take antibiotic | 82 (28%) | 21 (24%) | 61 (30%) |  |
| Noravirus genotype |  |  |  | 0.162 |
| GI infection | 38 (16%) | 17 (20%) | 21 (13%) |  |
| GII infection | 203 (84%) | 66 (80%) | 137 (87%) |  |
| Norovirus GI Ct-value, median (IQR) | 29 (24, 33) | 28 (24, 32) | 30 (24, 34) | 0.25 |
| Norovirus GII Ct-value, median (IQR) | 25 (21, 30) | 25 (21, 31) | 25 (21, 30) | 0.92 |
| Outcome at 15 day follow up |  |  |  | 0.59 |
| Died | 1 (<1%) | 0 (0%) | 1 (<1%) |  |
| Doing well at home | 278 (95%) | 84 (95%) | 194 (95%) |  |
| Recurrence of diarrhoea at home | 11 (4%) | 4 (5%) | 7 (3%) |  |
| Unknown | 3 (1%) | 0 (0%) | 3 (1%) |  |
| Acute gastroenteritis |  |  |  | 0.25 |
| No (main study controls) | 182 (62%) | 59 (67%) | 123 (60%) |  |
| Yes (main study cases) | 111 (38%) | 29 (33%) | 82 (40%) |  |
| NOTE |  |  |  |  |
| One patient with both GI and GII norovirus infection contribute to both groups in calculations.<br>The vesikari score for 89 cases without measured temperature was based on a score of 1 if they did not report fever and a score of 2 if they reported fever.<br>Vesikari score is adapted from: Ruuska, T., & Vesikari, T. (1990). Rotavirus Disease in Finnish Children: Use of Numerical Scores for Clinical Severity of Diarrhoeal Episodes. Scandinavian Journal of Infectious Diseases, 22(3), 259–267 |  |  |  |  |

<sup>1</sup>126 norovirus negative AGE cases were randomly selected to be tested for other pathogens.

Among GII noroviruses, overlapping Ct-value distributions were observed between norovirus-only and norovirus co-detection AGE cases (Figure 3A), as well as between norovirus-only AGE cases and norovirus-positive non-AGE controls (with either norovirus-only or norovirus co-detections) (Figure 3B, 3C). The Ct-value distributions for GI noroviruses seemed to indicate higher viral loads among norovirus-only AGE cases compared with norovirus co-detection AGE cases (Figure 3D), although the sample size was small. Restricting these analyses to children aged <2 years showed similar results (Supplementary Figure 1).

**Figure 3:**
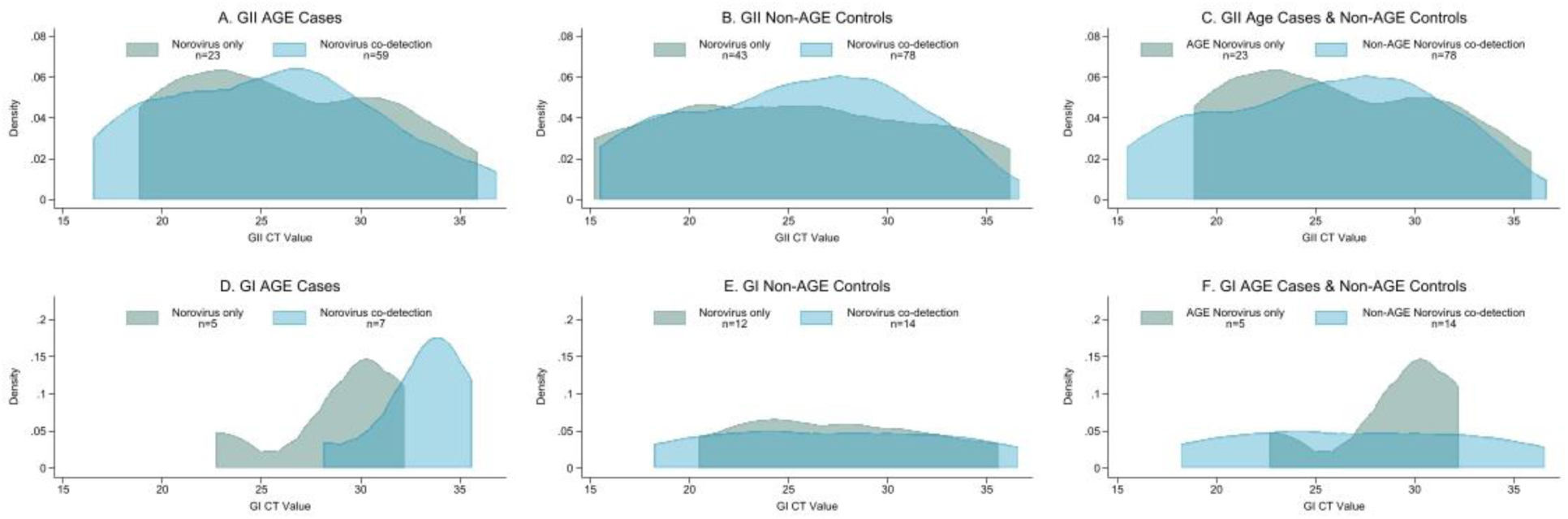
Distribution of GI and GII norovirus cycle threshold (ct) values among hospitalized norovirus-positive acute gastroenteritis (AGE) cases and non-AGE controls. Includes patients of all ages (N=240) from 10 tertiary care hospitals in Bangladesh from March 2018 to October 2021.

Higher positivity rates of norovirus and rotavirus were observed between November and April. However, the overall prevalence of rotavirus during our study period was much higher compared to that of norovirus (61% versus 12%, respectively) (Figure 4).

**Figure 4:**
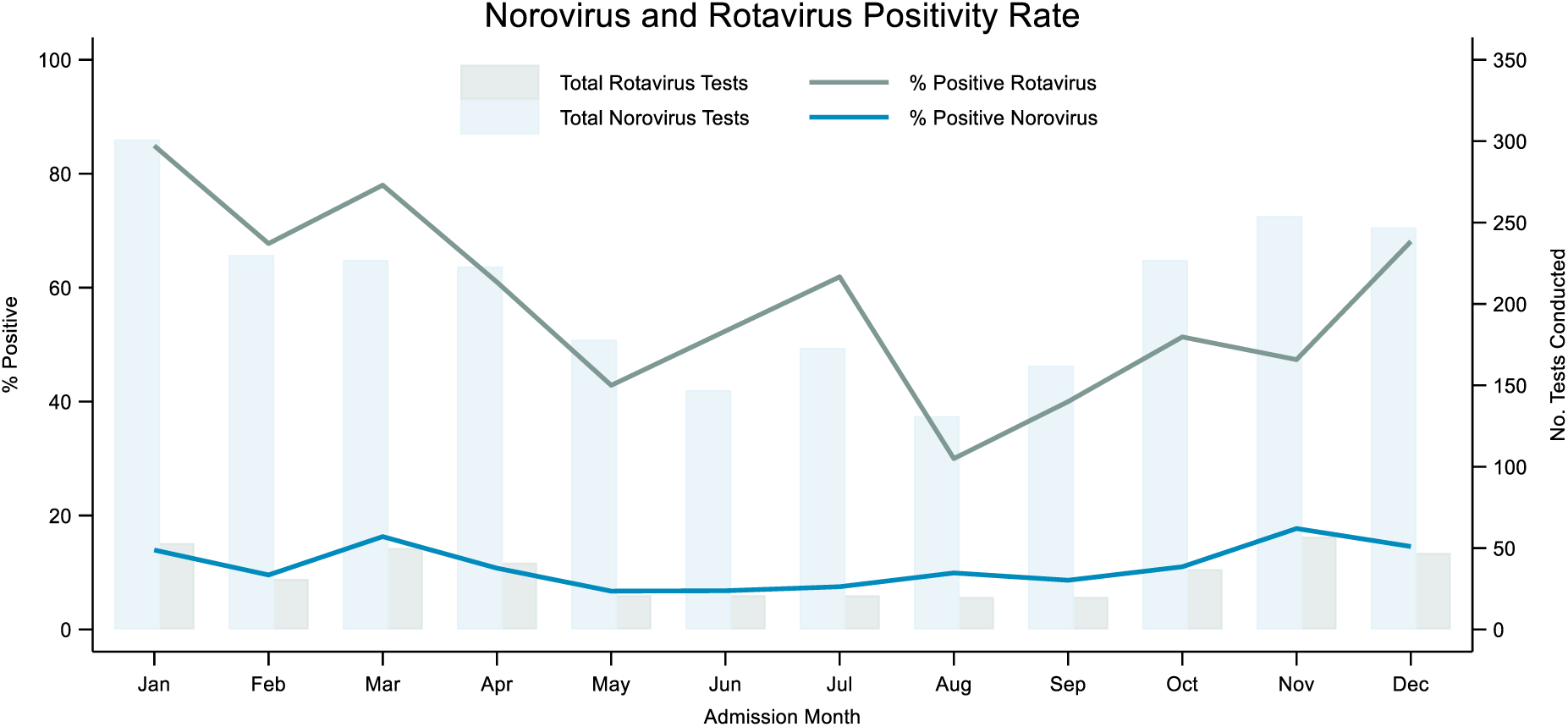
Monthly norovirus and rotavirus positivity rate among patients enrolled in 10 tertiary care hospitals in Bangladesh from March 2018 to October 2021. Monthly positivity rate is calculated as the sum of positive stool specimens at month of admission across all years divided by the sum of patients tested during that month across all years. All 2,500 enrolled AGE cases and non-AGE controls were tested for norovirus, but only a subset of 293 AGE cases and non-AGE controls who tested positive for norovirus and 126 randomly selected AGE cases who tested negative for norovirus were tested for rotavirus. As such, the rotavirus positivity rate is not reflective of the full study sample. Based on these data, the overall prevalence for norovirus and rotavirus across the study period was 12% (293/2,500) and 61% (255/419), respectively.

## Discussion

Our data underscore the high diversity of enteric pathogens in Bangladesh and the primacy of viruses among circulating enteropathogens. Mixed detections with norovirus and an additional enteric pathogen were extremely common among AGE cases (this was evident in almost 75% of norovirus detections), highlighting the difficulty in attributing diarrhea specifically to norovirus in Bangladesh. Among norovirus-positive stool samples with at least one other pathogen, 86% contained an additional viral pathogen, with norovirus and rotavirus co-detections predominating in 68% of these specimens. This high proportion of co-detections, primarily of norovirus and rotavirus, aligns with findings from others in Bangladesh.[17–20] Also, as in prior studies[21, 22], viral infections were more common in infants and children <5 years of age, while bacterial pathogens were more common among older children and adults. Even though viral pathogens predominated among patients with diarrhea, 70% of patients with AGE received antibiotics prior or during their hospital stay, emphasizing the need for more judicious use of antibiotics in Bangladesh to help minimize antimicrobial resistance and unnecessary medical costs,[23] particularly among infants and young children who are more likely to be infected with an enteric virus.

We also show that comprehensive testing of stool specimens can identify an enteropathogen in most AGE patients in a highly endemic setting. Extrapolating our detection rates to untested specimens, we estimated that 86% of hospitalized patients with AGE would have had one or more enteric pathogens detected with complete testing, similar to what has been reported in prior studies in Bangladesh, where an enteric virus or bacteria was found in 75%– 82% of patients with diarrhea.[4, 21, 22] Yet, differences in the strength of association between pathogens and clinical disease limit diagnostic interpretability, particularly when using sensitive molecular assays.[24]

Data are limited regarding whether co-infections with multiple enteric pathogens may be associated with more severe clinical outcomes compared with single-pathogen infections. While a few studies report more overt symptoms and dehydration with enteropathogen co-infections [25], most others have found no correlation between symptoms or the length of hospital stay between single and mixed infections.[17–19, 26, 27] Among AGE cases in our study, we did not observe differences in disease severity between norovirus co-detections compared to norovirus-only detections. Moreover, at first instance, the prevalence ratio of norovirus co-detections among AGE cases (7% [82/1250]) and non-AGE controls (10% [123/1250]) remained below one, suggesting limited association between norovirus co-detections and diarrhea symptoms. Nevertheless, our findings on co-detections were limited to norovirus-positive patients only, and we did not compare disease severity for other pathogen combinations, so these could be examined further. Of note, at least one other study suggests synergistic pathogenicity between rotavirus and certain co-infecting pathogens (e.g., *Giardia lamblia*) in causing diarrhea.[29]

In settings where gastroenteritis pathogens are commonly detected among asymptomatic individuals, quantification of pathogens in stool specimens from symptomatic patients and asymptomatic controls has been used to better characterize the etiologies of AGE, with higher viral loads commonly found in cases compared to controls.[24, 30, 31] In Bangladesh, we found largely overlapping norovirus Ct-values among hospitalized AGE cases and non-AGE controls, suggesting that any etiologic attribution to norovirus using Ct-values would be small.[32] We explored whether the presence of co-infections could help disentangle the relationship between viral load and symptomatic infection. We expected lower Ct-values (higher viral loads) among AGE cases with norovirus-only detections, in which norovirus was the only identified cause of the symptoms, compared to AGE cases with norovirus co-detections, in which at least one other alternative cause for the presenting symptoms was detected. Furthermore, we expected this difference to be more prominent when comparing AGE cases with norovirus-only detections versus non-AGE controls (either norovirus-only or norovirus co-detections), who had no AGE symptoms, and in which norovirus was likely an incidental finding. In both comparisons, Ct-values largely overlapped among groups, suggesting potential limitations in using viral load independently to estimate the burden of norovirus in specific settings.

As in prior studies,[2, 33], we document the very high burden of rotavirus-associated AGE hospitalizations in Bangladesh. The prevalence of rotavirus among AGE cases (69%) far exceeded that of norovirus (9%), and rotavirus positivity rates surpassed 80% during months of peak rotavirus activity. Our data reiterate the considerable benefits a rotavirus vaccine program could have in Bangladesh, by directly reducing rotavirus hospitalizations and their associated costs,[34] and indirectly by increasing the availability of hospital beds for use by children with other illnesses.[35] Although not assessed in this study, the higher burden (and pathogenicity) of rotavirus compared to norovirus could have influenced our prior findings of higher rates of norovirus among non-AGE controls compared to AGE cases.[5] In the presence of a co-circulating pathogen with a greater burden and pathogenicity than norovirus, then the denominator of diarrhea cases could be inflated, and thus the positivity of norovirus among diarrhea cases would be lowered. In contrast, the prevalence of this pathogen would be expected to be low among non-diarrhea controls and thus have a negligible effect on the positivity of norovirus among non-diarrhea controls. Additional data are needed to evaluate this hypothesis.

Several limitations should be considered. Testing to detect other enteropathogens was limited to specimens from AGE cases and non-AGE controls who tested positive for norovirus, preventing measurement of the true prevalence of these enteropathogens. However, we tested a random sample of AGE cases who tested negative for norovirus to get a sense of the overall prevalence of these enteropathogens among AGE cases. Over two-thirds of our study sample received antibiotics before or during hospitalization, which could have decreased the yield of bacterial pathogens, although we used molecular assays and not cultures for the detection of bacteria. On the other hand, the use of molecular assays could have led to an overestimation of pathogen burden in both AGE cases and non-AGE controls. An appreciation of any differences in disease severity in AGE cases with norovirus-only versus norovirus co-detections was limited because all AGE cases were hospitalized, so their illnesses reflect the higher end of the severity spectrum. In addition, we did not consider the implications of other comorbidities (such as malnutrition) that might affect disease severity. A high proportion (∼60%) of non-AGE controls had stool specimens collected three or more days after admission,[5] and delays in specimen collection are associated with higher Ct-values.[31] However, the expectation of higher Ct-values among non-AGE controls than those seen among AGE cases was not observed. Late collection of stool specimens also allows for the possibility that any positive non-AGE control could be due to nosocomial transmission, and such recent hospital-acquired infections could have biased Ct-values in the opposite direction (towards lower Ct-values). However, we observed no differences in the distribution of Ct-values among non-AGE controls with stool specimens collected within and after 2 days from admission (Supplementary Figure 2).

In summary, our study highlights the vast diversity of diarrheal pathogens co-circulating in Bangladesh. The high predominance of viral enteropathogens, mostly rotavirus, underscores the need for more judicious use of antibiotics among diarrheal patients, and highlights the importance of introducing the rotavirus vaccine in Bangladesh. We underline a few key issues impeding a complete assessment of the role of norovirus in diarrhea in Bangladesh: the high proportion of asymptomatic carriage of noroviruses, the frequent detection of other enteropathogens in combination with norovirus among diarrhea cases, and the overlapping Ct-value distributions in patients with and without diarrhea, even when comparing Ct-values between AGE cases in which only norovirus was identified and non-AGE controls. New approaches are needed to improve our understanding of the contribution of different enteropathogens to diarrhea in high-burden settings,[32] as well as how the prevalence of noroviruses in cases and controls might be affected by the presence of other co-circulating enteropathogens.

## Data Availability

All data produced in the present study are available upon reasonable request to the authors. The copyright of the data solely lies with affiliated organizations.

## Acknowledgement

We want to thank all study participants for their time. The International Center for Diarrheal Disease Research (ICDDR,B) acknowledges the commitment of the US Centers for Disease Control and Prevention, Atlanta, Georgia, to its research efforts. ICDDR,B is also grateful to the Governments of Bangladesh and Canada for core/unrestricted support.

