## Supplementary table 1 for "High Levels of Co-detection of Norovirus and Other Enteric Pathogens in Hospitalized Patients with and without Acute Gastroenteritis in Bangladesh"

**Supplementary Table 1.** Characteristics of patients hospitalized in ten tertiary hospitals in Bangladesh from March 2018 to October 2021 by acute gastroenteritis (AGE) cases and non-AGE controls and type of norovirus infection

|  |  | **AGE Cases** | | **Non-AGE Controls** | |
| --- | --- | --- | --- | --- | --- |
|  | **Total** | **Norovirus only-detection** | **Norovirus co-detection** | **Norovirus only-detection** | **Norovirus co-detection** |
|  | N=293 | N=29 | N=82 | N=59 | N=123 |
| **Demographic and Socioeconomic** |  |  |  |  |  |
| Division |  |  |  |  |  |
| Barishal | 61 (21%) | 3 (10%) | 15 (18%) | 9 (15%) | 34 (28%) |
| Chattogram | 26 (9%) | 1 (3%) | 5 (6%) | 5 (8%) | 15 (12%) |
| Dhaka | 37 (13%) | 2 (7%) | 13 (16%) | 8 (14%) | 14 (11%) |
| Khulna | 20 (7%) | 1 (3%) | 5 (6%) | 5 (8%) | 9 (7%) |
| Rajshahi | 110 (38%) | 16 (55%) | 32 (39%) | 25 (42%) | 37 (30%) |
| Rangpur | 16 (5%) | 4 (14%) | 3 (4%) | 5 (8%) | 4 (3%) |
| Sylhet | 23 (8%) | 2 (7%) | 9 (11%) | 2 (3%) | 10 (8%) |
| Age (years), median (IQR) | 1 (0.6, 2) | 1 (0.8, 20) | 1 (0.7, 1) | 1 (0.5, 2) | 1 (0.5, 3) |
| Age category (years) |  |  |  |  |  |
| 0-4 | 245 (88%) | 20 (74%) | 71 (90%) | 49 (91%) | 105 (88%) |
| 5-17 | 6 (2%) | 0 (0%) | 1 (1%) | 3 (6%) | 2 (2%) |
| 18+ | 28 (10%) | 7 (26%) | 7 (9%) | 2 (4%) | 12 (10%) |
| Sex |  |  |  |  |  |
| Female | 91 (31%) | 10 (34%) | 23 (28%) | 20 (34%) | 38 (31%) |
| Male | 202 (69%) | 19 (66%) | 59 (72%) | 39 (66%) | 85 (69%) |
| Number of permanent household members, median (IQR) | 5 (4, 6) | 5 (4, 6) | 5 (4, 6) | 5 (4, 6) | 5 (4, 6) |
| Maternal education status for patients <18 years of age |  |  |  |  |  |
| Illiterate | 11 (4%) | 0 (0%) | 3 (4%) | 4 (7%) | 4 (4%) |
| Primary/PCS | 75 (29%) | 6 (30%) | 21 (29%) | 14 (25%) | 34 (31%) |
| Class eight/JSC | 94 (37%) | 7 (35%) | 24 (33%) | 21 (38%) | 42 (39%) |
| SSC/HSC | 63 (25%) | 5 (25%) | 23 (32%) | 14 (25%) | 21 (19%) |
| Graduate/Master and above | 9 (4%) | 2 (10%) | 2 (3%) | 2 (4%) | 3 (3%) |
| Religous schooling | 4 (2%) | 0 (0%) | 0 (0%) | 0 (0%) | 4 (4%) |
| Monthly family income (taka), median (IQR) | 15,000 (10000, 20000) | 15,000 (7000, 20000) | 15,000 (10000, 22000) | 12,000 (9000, 25000) | 14,500 (10000, 20000) |
| Shared bathroom or toilet with another household |  |  |  |  |  |
| No | 190 (65%) | 24 (83%) | 56 (68%) | 32 (54%) | 78 (64%) |
| Yes | 102 (35%) | 5 (17%) | 26 (32%) | 27 (46%) | 44 (36%) |
| **Clinical** |  |  |  |  |  |
| Patient transferred from another facility/hospital |  |  |  |  |  |
| No | 279 (95%) | 27 (93%) | 70 (85%) | 59 (100%) | 123 (100%) |
| Yes | 14 (5%) | 2 (7%) | 12 (15%) | 0 (0%) | 0 (0%) |
| Contact in last 10 days before illness with someone with diarrhoea or vomitting |  |  |  |  |  |
| No | 264 (90%) | 25 (86%) | 70 (85%) | 55 (93%) | 144 (93%) |
| Yes | 28 (10%) | 4 (14%) | 12 (15%) | 4 (7%) | 8 (7%) |
| Duration of hospital stay (days), median (IQR) | 3 (2, 7) | 3 (1, 3) | 2 (1, 3) | 5 (3, 10) | 6 (3, 8) |
| Vesikari score |  |  |  |  |  |
| Moderate (7-10) | 26 (23%) | 6 (21%) | 17 (21%) | - | - |
| Severe (>=11) | 85 (77%) | 23 (79%) | 65 (79%) | - | - |
| Antibiotic use before or during hospitalization prior to stool collection |  |  |  |  |  |
| No | 82 (28%) | 11 (38%) | 22 (27%) | 10 (17%) | 39 (32%) |
| Yes | 211 (72%) | 18 (62%) | 60 (73%) | 49 (83%) | 84 (68%) |
| Antibiotic usage |  |  |  |  |  |
| Oral | 12 (4%) | 2 (7%) | 10 (12%) | 0 (0%) | 0 (0%) |
| Injectable | 2 (1%) | 2 (7%) | 0 (0%) | 0 (0%) | 0 (0%) |
| Given at hospital | 197 (67%) | 14 (48%) | 50 (61%) | 49 (83%) | 84 (68%) |
| Did not take antibiotic | 82 (28%) | 11 (38%) | 22 (27%) | 10 (17%) | 39 (32%) |
| Noravirus Genotype |  |  |  |  |  |
| GI | 38 (16%) | 5 (18%) | 7 (11%) | 12 (22%) | 14 (15%) |
| GII | 203 (84%) | 23 (82%) | 59 (89%) | 43 (78%) | 78 (85%) |
| Norovirus GI Ct-value, median (IQR) | 29 (24, 33) | 30 (28, 31) | 34 (32, 35) | 27 (23, 32) | 27 (22, 33) |
| Norovirus GII Ct-value, median (IQR) | 25 (21, 30) | 26 (22, 31) | 25 (20, 29) | 25 (19, 31) | 26 (21, 31) |
| Outcome at 15 day follow up |  |  |  |  |  |
| Died | 1 (<1%) | 0 (0%) | 0 (0%) | 0 (0%) | 1 (1%) |
| Doing well at home | 278 (95%) | 29 (100%) | 80 (98%) | 55 (93%) | 114 (93%) |
| Recurrence of diarrhoea, at home | 11 (4%) | 0 (0%) | 0 (0%) | 4 (7%) | 7 (6%) |
| Unknown | 3 (1%) | 0 (0%) | 2 (2%) | 0 (0%) | 1 (1%) |
| Acute gastroenteritis |  |  |  |  |  |
| No (main study controls) | 182 (62%) | - | - | 59 (100%) | 123 (100%) |
| Yes (main study cases) | 111 (38%) | 29 (100%) | 82 (100%) | - | - |
| NOTE |  |  |  |  |  |
| Numbers may not add to column totals due to missing.  One norovirus only infected non-AGE control has both GI and GII norovirus infection and contributes to both groups in calculations.  The vesikari score for 89 cases without measured temperature was based on a score of 1 if they did not report fever and a score of 2 if they reported fever | | | | | |
