## Supplementary Table 2a for "High Levels of Co-detection of Norovirus and Other Enteric Pathogens in Hospitalized Patients with and without Acute Gastroenteritis in Bangladesh"

**Supplementary Table 2a.** Distribution of enteric pathogens identified among randomly selected norovirus negative hospitalized acute gastroenteritis (AGE) cases in Bangladesh. N=126

|  | **Norovirus Negative AGE Cases** |
| --- | --- |
|  | N=126 |
| *No other pathogens detected* | 19 (15%) |
| *One or more other pathogens detected* | 107 (85%) |
| **Subset of one or more other pathogens detected:** | **N=107** |
| *One Pathogen Detected* | 68 (54%) |
| Virus: Rotavirus | 65 (96%) |
| Parasite: Giardia lamblia | 1 (1%) |
| Bacteria: Campylobacter jejuni | 1 (1%) |
| Virus: Adenovirus | 1 (1%) |
| *Two Pathogens Detected* | 33 (26%) |
| Virus - Virus: Rotavirus + Adenovirus | 15 (45%) |
| Virus - Bacteria: Rotavirus + Shigella | 6 (18%) |
| Virus - Virus: Rotavirus + Astrovirus | 4 (12%) |
| Virus - Parasite: Rotavirus + Cryptosporidium | 2 (6%) |
| Virus - Bacteria: Rotavirus + Campylobacter jejuni | 2 (6%) |
| Virus - Bacteria: Rotavirus + Salmonella | 1 (3%) |
| Virus - Bacteria: Adenovirus + Shigella | 1 (3%) |
| Virus - Bacteria: Adenovirus + Campylobacter jejuni | 1 (3%) |
| Virus - Parasite: Rotavirus + Giardia lamblia | 1 (3%) |
| *Three Pathogens Detected* | 6 (5%) |
| Virus - Virus - Bacteria: Rotavirus + Adenovirus + Campylobacter jejuni | 2 (33%) |
| Virus - Bacteria - Parasite: Adenovirus + Campylobacter jejuni + Cryptosporidium | 1 (17%) |
| Virus - Bacteria - Parasite: Rotavirus + Shigella + Giardia lamblia | 1 (17%) |
| Virus - Bacteria - Bacteria: Rotavirus + Campylobacter jejuni + Shigella | 1 (17%) |
| Virus - Virus - Bacteria: Rotavirus + Adenovirus + Shigella | 1 (17%) |
| NOTE: |  |
| Column percentages presented. Totals for each pathogen combination sum to respective group header. | |
| Other pathogens tested for include viruses (rotavirus, adenovirus, astrovirus, sapovirus), bacteria (*Clostridium difficile, Salmonella*, *Shigella, Campylobacter jejuni, Yersinia enterocolitica*, enteroinvasive *Escherichia coli*), and parasites (*Cryptosporidium*, *Entamoeba histolytica*, *Giardia lamblia*). | |
