## Supplementary Table 2b for "High Levels of Co-detection of Norovirus and Other Enteric Pathogens in Hospitalized Patients with and without Acute Gastroenteritis in Bangladesh"

**Supplementary Table 2b.** Distribution of enteric pathogens identified among randomly selected norovirus negative hospitalized acute gastroenteritis (AGE) cases in Bangladesh by age group. N=126

|  | **Age Group (years)** | | | **All** |
| --- | --- | --- | --- | --- |
|  | **0-4** | **5-17** | **18+** |  |
|  | N=44 | N=41 | N=41 | N=126 |
| Other pathogens detected |  |  |  |  |
| One or more | 42 (95%) | 35 (85%) | 30 (73%) | 107 (85%) |
| None | 2 (5%) | 6 (15%) | 11 (27%) | 19 (15%) |
| Any viral pathogens detected |  |  |  |  |
| Yes | 42 (95%) | 33 (80%) | 30 (73%) | 105 (83%) |
| No | 2 (5%) | 8 (20%) | 11 (27%) | 21 (17%) |
| Any bacterial pathogens detected |  |  |  |  |
| Yes | 4 (9%) | 10 (24%) | 4 (10%) | 18 (14%) |
| No | 40 (91%) | 31 (76%) | 37 (90%) | 108 (86%) |
| Any parasitic pathogens detected |  |  |  |  |
| Yes | 3 (7%) | 3 (7%) | 0 (0%) | 6 (5%) |
| No | 41 (93%) | 38 (93%) | 41 (100%) | 120 (95%) |
| Column percentages. |  |  |  |  |
| Other pathogens tested for include: 1) Viruses (rotavirus, adenovirus, astrovirus, sapovirus), 2) Bacteria (*Clostridium difficile, Salmonella*, *Shigella, Campylobacter jejuni, Yersinia enterocolitica*, enteroinvasive *Escherichia coli*), and 3) Parasites (*Cryptosporidium*, *Entamoeba histolytica*, *Giardia lamblia*). | | | | |
