## Supplementary Figure 1 for "High Levels of Co-detection of Norovirus and Other Enteric Pathogens in Hospitalized Patients with and without Acute Gastroenteritis in Bangladesh"

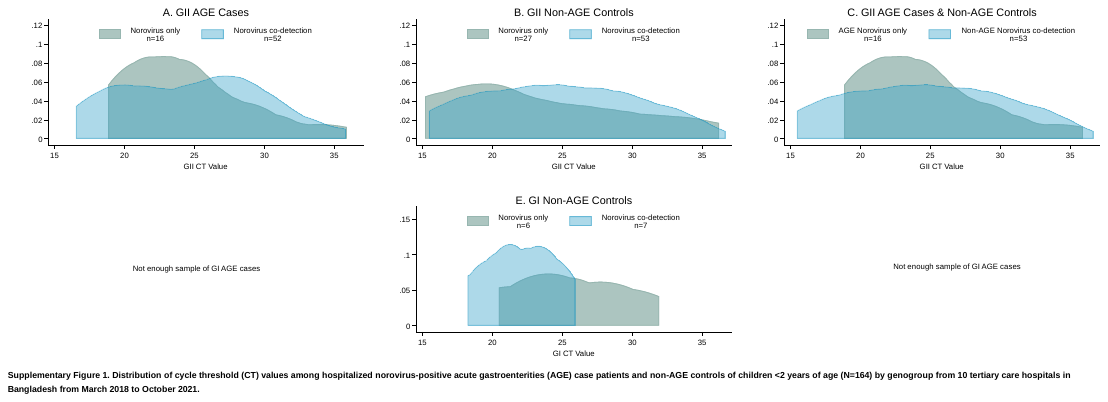

**Supplementary Figure 1:** Distribution of cycle threshold values (Ct-values) among hospitalized acute gastroenteritis (AGE) cases and non-AGE controls of children < 2 years of age (N=164) by genogroup from 10 tertiary care hospitals in Bangladesh from March 2018 to October 2021
