## Supplementary Figure 2 for "High Levels of Co-detection of Norovirus and Other Enteric Pathogens in Hospitalized Patients with and without Acute Gastroenteritis in Bangladesh"

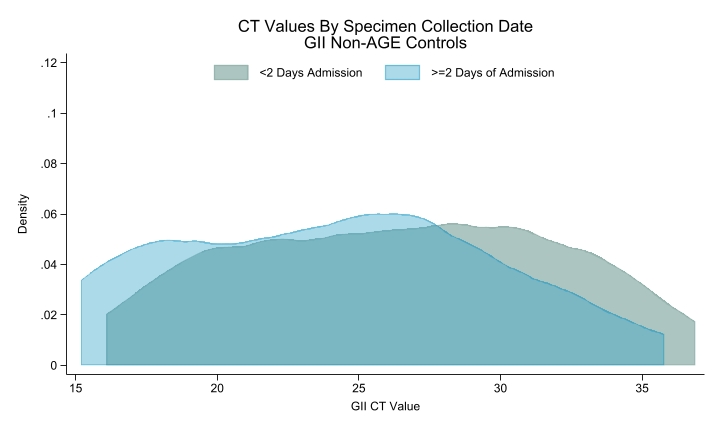

**Supplementary Figure 2.** Distribution of Ct-values of GII non-AGE controls by specimen collection date.
